# Accurate and Rapid Molecular Subgrouping of High-Grade Glioma via Deep-learning-assisted Label-free Fiberoptic Raman Spectroscopy

**DOI:** 10.1101/2023.07.03.23292176

**Authors:** Chang Liu, Jiejun Wang, Jianghao Shen, Xun Chen, Nan Ji, Shuhua Yue

**Affiliations:** Key Laboratory of Biomechanics and Mechanobiology (Beihang University), Ministry of Education, Institute of Medical Photonics, Beijing Advanced Innovation Center for Biomedical Engineering, School of Biological Science and Medical Engineering, Beihang University, Beijing 100191, China; Department of Neurosurgery, Beijing Tiantan Hospital, Capital Medical University, Beijing 100050, China; School of Engineering Medicine, Beihang University, Beijing 100191, China

## Abstract

Glioma are often impossible to visualize discrimination within different grades and staging, especially for glioma molecular subgrouping which is highly related with surgery strategy and prognosis. Based on glioma guideline published on 2021, molecular subgroups such as IDH, 1p/19q etc. need to be detected to classify the subgroups (astrocytoma, oligodendroglioma, GBM) from high-grade glioma and guide the personalized treatment. However, timely intraoperative technology is limited to identify molecular subgroups of glioma tissues. To address this problem, we develop a deep learning-guided fiberoptic Raman diagnostic platform to assess its ability of real-time high-grade glioma molecular subgrouping. The robust Raman diagnostic platform is established using convolutional neural networks (ResNet) together with fingerprint spectra acquired within 3 seconds. We have acquired a total of 2358 Raman spectra from 743 tissue sites (astrocytoma: 151; oligodendroglioma:150; GBM: 442) of 44 high-grade glioma patients (anaplastic astrocytoma: 7; anaplastic oligodendroglioma:8; GBM: 29). The optimized ResNet model provides an overall mean diagnostic accuracy of 84.1% (sensitivity of 87.1% and specificity of 81.5%) for identifying 7 molecular subgroups (e.g., IDH, 1p/19q, MGMT, TERT, EGFR, Chromosome 7/10, CDKN2A/B) of high-grade glioma, which is superior to the best diagnosis performance using PCA-SVM and UMAP. We further investigate the saliency map of the best ResNet models using the correctly predicted Raman spectra. The specific Raman features that are related to the tumor-associated biomolecules (e.g., collagens, and lipids) validate the robustness of ResNet diagnostic model. This potential intraoperative technology may therefore be able to diagnosis molecular subgroups of high-grade glioma in real time, making it an ideal guide for surgical resection and instant post-operative decision-making.

## Introduction

High-grade gliomas are the most common and aggressive primary tumors of the central nervous system^1^. The molecular genetic stratification of gliomas is important as more evidence emerges of the predictive and prognostic implications of different genetic subgroups^2^.As an major treatment, gross total resection is demonstrated to improve patient’s progression-free and overall survival. Moreover, molecular subgrouping techniques, which can provide predictive and prognostic information, are introduced into the surgical workflow This approach would allow the surgeon to optimized their extent of surgical resection for different gene profiles^3–5^.

Based on the latest glioma grading guideline of World Health Organization (WHO) published on June 2021 at the time of writing^6^, high-grade glioma include three subgroups, including grade 3 to 4 astrocytoma, grade 3 oligodendroglioma, and grade 4 glioblastoma (GBM). Based on the histological glioma type and molecular subgroups together (e.g. IDH, 1p/19q, MGMT, TERT, EGFR, Chromosome7/10, CDKN2A/B), high-grade glioma could be classified into three types. For instance, GBMs, which are considered to be with high malignancy and poor prognosis, are discriminated from the other two high-grade glioma subgroup by IDH wild subgrouping with histological GBM type.^7^. Glioma without histological features is also diagnosed to be GBM with IDH wild and either EGFR amplification, TERT mutation or chromosome copynumber +7/-10 (combined entire chromosome 7 gain/ entire chromosome 10 loss)^6^. Oligodendroglioma is currently defined on histological feature with1p/19q codeletion, and IDH mutation^8^. Astrocytoma is defined on histological feature with 1p/19q intact, and IDH wild^9^. Patients with 1p/19q codeletion have better prognosis and are sensitive to chemotherapy and radiotherapy.

The current gold standard of the molecular subgrouping is immunohistochemistry (IHC), cytogenetic testing and next-generation sequencing^10–12^. However, next-generation sequencing is complex and time-consuming (1-2 weeks), which cannot provide molecular subgroups during surgery. To detect above molecular subgroups noninvasively, rapidly, and accurately, magnetic resonance imaging (MRI) shows the potential incorporating with machine learning algorithms. MRI diagnosed IDH subgrouping with an AUC of 0.888^13^, 1p/19q with an AUC of 0.811^14^, and MGMT subgrouping with an AUC of 0.898^15^. However, molecular subgroups (TERT, EGFR, Chromosome 7/10, and CDKN2A/B) have not been detected by MRI^16,17^. There is an urgent clinical need for comprehensive simultaneous detection of multiple molecular types. MRI also cannot uncover the rational biological explanation about IDH, 1p/19q, MGMT etc. subgroups simultaneously with robust diagnostic accuracy^18^. Therefore, rapid and non-destructive methods with higher molecular selectivity are required.

Raman spectroscopy is a label-free optical vibrational spectroscopy technique, which can investigate specific biomolecular compositions of tissues, and has been explored for diagnosis of multiple human cancers^19–26^.

Previous studies demonstrated that Raman spectroscopy enables intraoperative brain cancer detection in humans^27,28^. Leblond et al. discriminated dense cancer from normal brain with a sensitivity of 93% and a specificity of 91%^19^. Galli et al. achieved to classify oligodendroglioma, astrocytoma and GBM with correct rates of 94%, 86% and 90%^29^. However, IDH1-mutant, and 1p/19q codeletion, were discerned only with a correct rate of 81%. Uckermann et al. classified IDH1 mutation with a correct rate of 89%^30^. Sciortino et al. studied the Raman spectral difference between IDH wild and mutation, which achieves an accuracy of 87%^31^. Livermore et al. also improved IDH subgrouping with the sensitivity and specificity of 91% and 95%^32^. However, other subgroups (TERT, EGFR, Chromosome 7/10, CDKN2/B) have not been investigated using the Raman biopsy technologies. Hollon et al. developed an artificial-intelligence-based molecular classification (IDH, 1p/19q and ATRX) of diffuse gliomas with the accuracy of 93.3 ± 1.6% using stimulated Raman histology (SRH)^33^. However, this approach scanning overall tissue (>1 cm) will cost too much time (>1 hour).

To fulfil the unmet clinical need, here we develop a molecular-specific convolutional neural network (ResNet) model together with fiberoptic Raman spectroscopy to achieve real-time and accurate diagnosis of molecular subgroups in high-grade glioma. We have acquired a total of 2358 Raman spectra from 743 tissue sites (astrocytoma: 151; oligodendroglioma:150; GBM: 442) of 44 high-grade glioma patients (astrocytoma: 7; oligodendroglioma:8; GBM: 29). Our spectral ResNet model provides an overall mean diagnostic accuracy of 84.1% (sensitivity of 87.1% and specificity of 81.5%) for identifying 7 molecular (IDH, 1p/19q, MGMT, TERT, EGFR, Chromosome 7/10, and CDKN2A/B) subgroups of high-grade glioma simultaneously. The AUCs of all the molecular subgrouping were larger than 0.8, and the AUCs of IDH, 1p/19q and CDKN were larger than 0.9. The accuracy together with the sensitivity and specificity of the model was calculated using an external testing dataset from extra samples, which avoids spectra from the same samples for train and test set. The fingerprint spectra acquisition for each tissue cite is within 3 seconds, and the diagnostic time-cost for 7 molecular subgrouping simultaneously using ResNet is 4 ms per spectra. The diagnostic performance of the deep learning model (ResNet) was then compared to that of the machine learning model (PCA-SVM) and manifold learning model (UMAP). We further investigated the saliency map with the ResNet to reveal the key biomolecular features for molecular subgrouping of high-grade glioma. It was found that the molecular weights of lipid and collagen in saliency map were relatively higher for high-grade glioma subgrouping. Together, the Raman technology could be integrated into the neurosurgical workflow for rapid and accurate identification of glioma molecular subgrouping with sensitivity beyond current capabilities.

## Results

### Workflow of high-grade glioma molecular subgrouping

As shown in **Figure 1**, the workflow of high-grade glioma and GBM molecular subgrouping by Raman spectroscopy was described. Based on 2021 WHO glioma classification guideline, we focus on 7 typical molecular subgroups (IDH, 1p/19q, MGMT, TERT, EGFR, Chromosome 7/10, CDKN2A/2B) of high-grade glioma. First, IDH and 1p/19q subgroups were identified to classify GBM from high-grade glioma. Then, molecular subgroups (MGMT methylation, TERT wildtype, EGFR amplification, CDKN homozygous deletion, Chromosome +7/-10) were identified in both high-grade glioma and GBM.

**Fig. 1.**
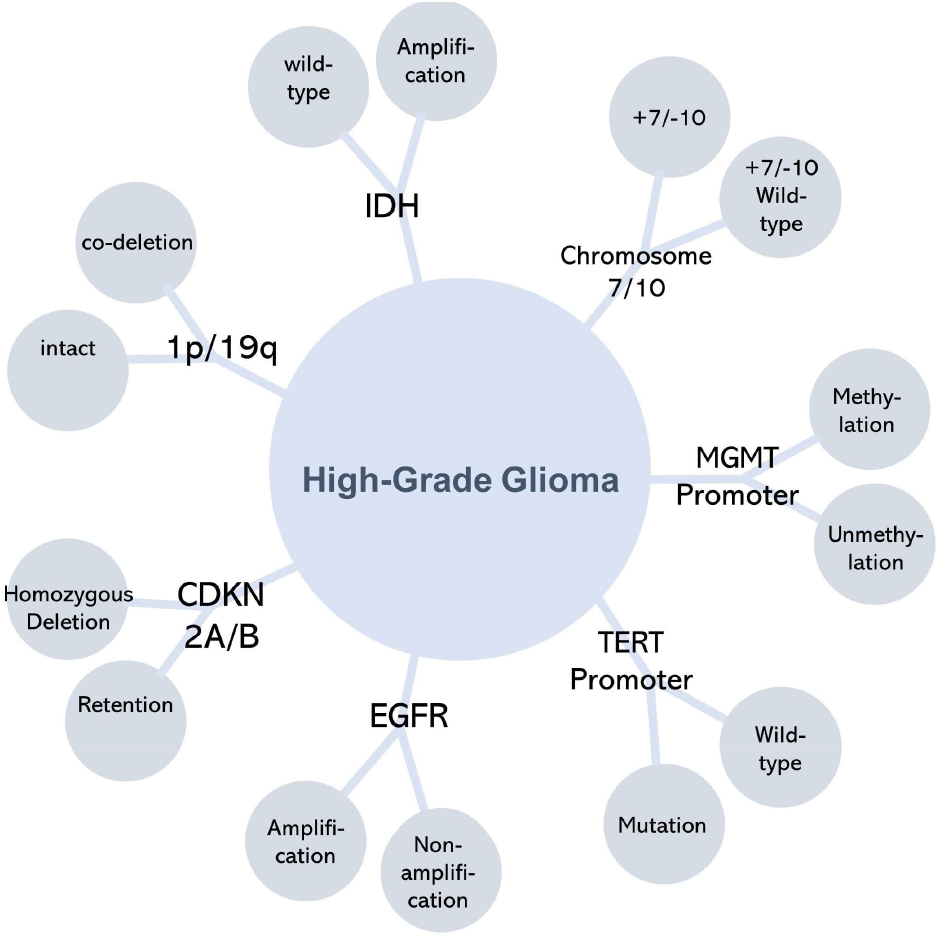
Workflow of high-grade glioma molecular subgrouping classification. According to 2021 WHO classification system, 7 typical molecular subgroups (IDH, 1p/19q, MGMT, TERT, EGFR, Chromosome7/10, CDKN2A/2B) were selected. First, high-grade glioma was classified for IDH and 1p/19q subgroups, and GBM was identified from high-grade glioma. Then, Raman of high-level glioma and GBM were classified for 7 molecular subgroups (MGMT methylation etc.).

As shown in **Figure 2a**, the workflow of deep-learning based Raman data classification and explanation analysis for high-grade glioma molecular subgrouping diagnosis was described. First, Raman spectra were acquired using fiberoptic Raman spectroscopy (**Supplementary Figure S1**) with 5 second for each tissue site (the details were described in the Method section). Second, the spectra have been preprocessed for auto-fluorescent removal, denoising, and min-max normalization. Third, for each molecular subgroup, the Raman data of the tissues was split into three datasets (80% for training, 10% for validation and 10% for test) for deep learning.

**Fig. 2.**
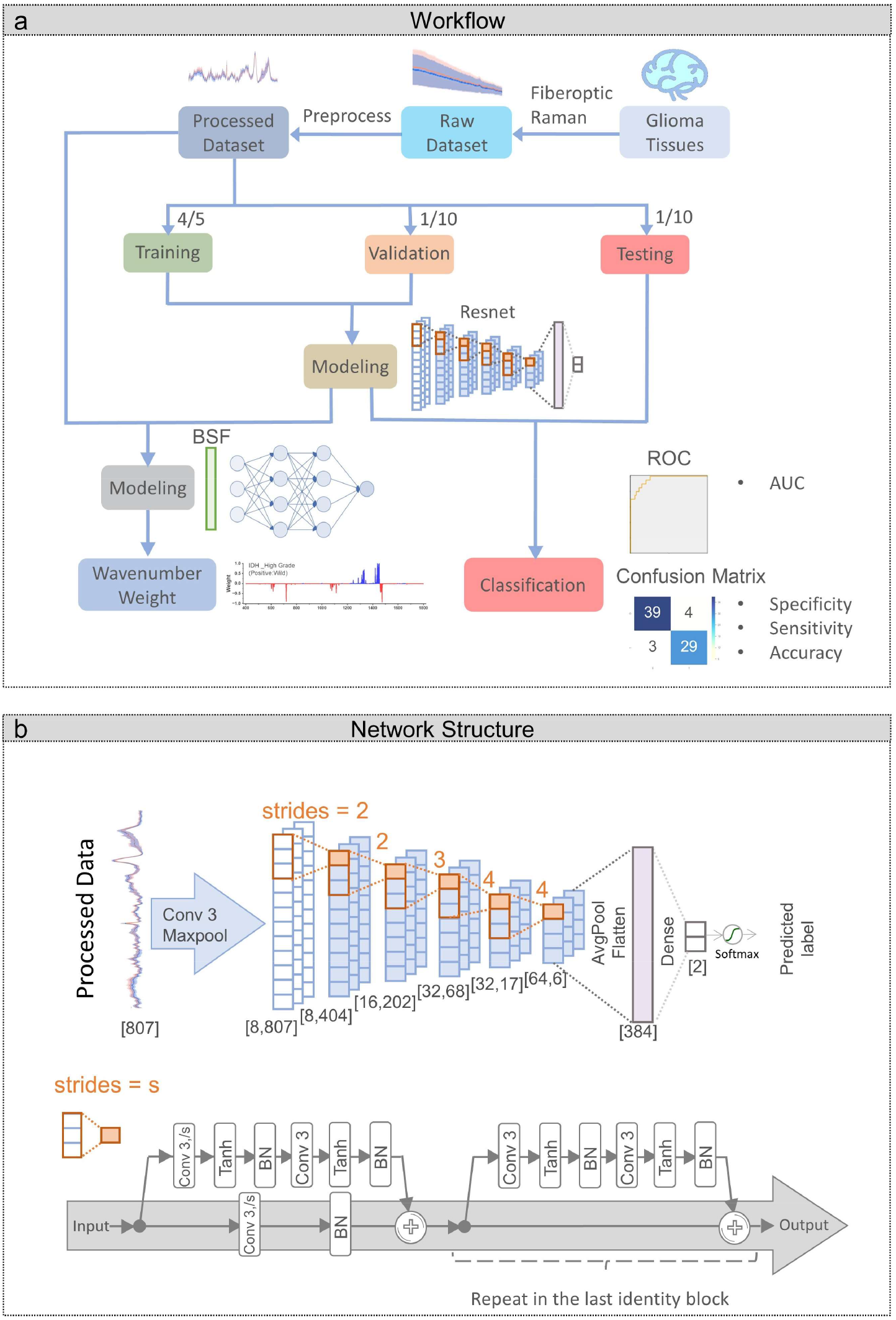
Workflow of each subgrouping classification. **a**, Workflow of each subgrouping modeling: the dataset preparing, Resnet model training for classification and BSF model training for weighing wavenumber contribution. **b**, Structure of the Resnet model designed. The numbers in bracket represent the size [(channels,) length] of the hidden layers. The orange numbers represent the strides of the first convolution layer in each identity block (orange dash line). Abbreviation: Conv: convolution layer with kernel size = 3 and strides = s, BN: batch normalization.

To avoid overfitting, the models were trained and validated during learning iteration. The prediction performance of the test set works as the final results of built subgrouping models. For model comparation, the same training sets were also put into the PCA−SVM and UMAP models for 10-fold cross-validation. To compare the performance between deep-learning and other models, the confusion matrix and the receiver operating characteristic (ROC) curve of each model were evaluated in the test set, as shown in **Figure 2a**.

Finally, in **Figure 2b**, we evaluated the significant biomolecular information used for diagnosis by saliency maps which indicate the contribution of different Raman wavenumber. The saliency maps were generated by difference spectrum and binary stochastic filtering (BSF)^34^. The detail of BSF restrained schematic is described in the method.

### Raman spectroscopy analysis of glioma tissues

Using our fiberoptic Raman spectroscopy, we acquired Raman data from 743 tissue sites of 44 high-grade glioma patients. **Figure 3** shows mean spectra from high-grade glioma patients with standard deviation. The spectra have been preprocessed with iterative multi-polynomial fitting, s-g filter and min-max normalization. Distinct tissue Raman peaks can be observed from all glioma (**Figure 3**) in the fingerprint region. For each molecular subgroup, difference spectrum was calculated by vector subtraction between mean spectra from positive and negative patients. In summary, spectra obtained are dominated by the CH2 scissoring deformations at 1440cm^-1^, near which the spectral difference is persistently observed. Peak of the bond are shifted in different molecular subgroups. Also pronounced are C=O stretching of protein backbone at 1661 cm^-1^, which is often referred to as amid I band. Amide III band between 1200 and 1380cm^-1^ is related to C-H and N-C bending deformations. The difference in Amide I and III bands is significant in 1p/19q (b), MGMT (c), TERT (d) and EGFR (e). Significant difference can also be observed near collagen bands at 854 and 938 cm^-1^ and lipid related signal from 492 to 604 cm^-1^.

**Fig. 3.**
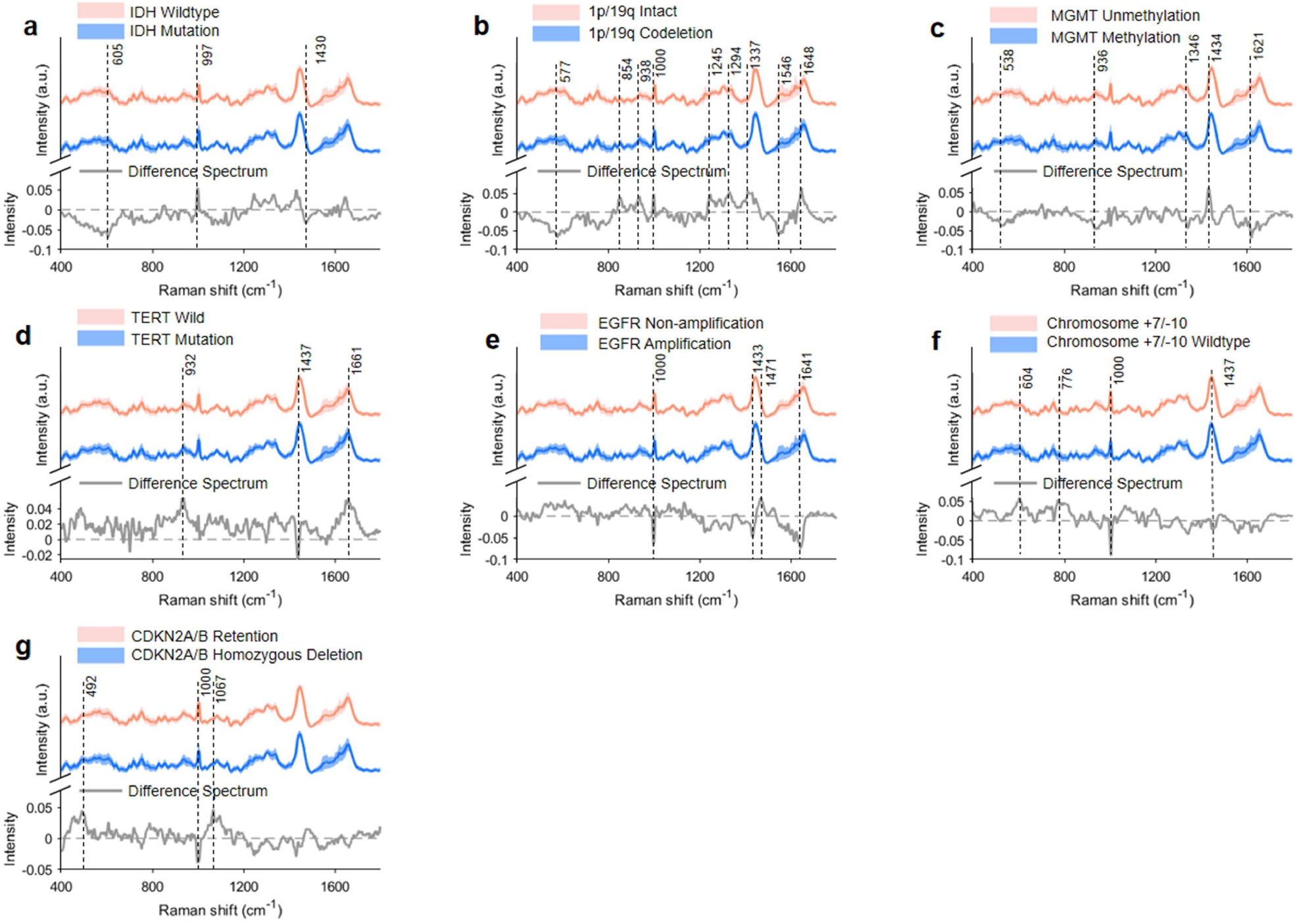
Mean FP Raman spectra of high-grade glioma molecular subgrouping for classification acquired from 743 tissue sites of 44 patients under Raman spectroscopy. **a**, Tissue Raman spectral differences between IDH wild and mutation (wild:442; mutation: 301). **b**,1p/19q intact and codeletion tissues (intact: 593; codeletion: 150). **c**, MGMT unmethylation and methylation tissues (methylation: 546; unmethylation: 197). **d**, TERT wildtype and mutation tissues (mutation: 508; wildtype: 235). **e**, EGFR non-amplification and amplification tissues (amplification: 241; non-amplification: 502). f, Gain of chromosome 7 & loss of chromosome 10 and wild-type (+7/-10: 192; wild-type: 551). g, CDKN2A/B homozygous deletion and retention (homozygous deletion: 226; retention: 517).

### Accurate and rapid glioma molecular subgrouping with deep learning

The models are first trained and optimized with both training dataset and validation dataset. Then the performance of each model is evaluated in the test dataset. **Figure 4** shows the ROCs of molecular identification from high-grade glioma. The AUCs of deep learning (ResNet) are larger than 0.95 in IDH, 1p/19q and CDKN2A/B. The AUCs in other subgroups also exceed 0.8, revealing the potential of Raman based subgrouping. In comparisons, **Figure 5** respectively show the sensitivity, specificity accuracy and AUC for high-grade glioma subgrouping. For the most two previously investigated subgroups (IDH and 1p19q), our deep learning models (Resnet) achieve overall diagnostic accuracies of 90.67% (sensitivity of 93.75% and specificity of 88.37%) and 86.67% (sensitivity of 90.00% and specificity of 85.45%) in the **table S2**.

**Fig. 4.**
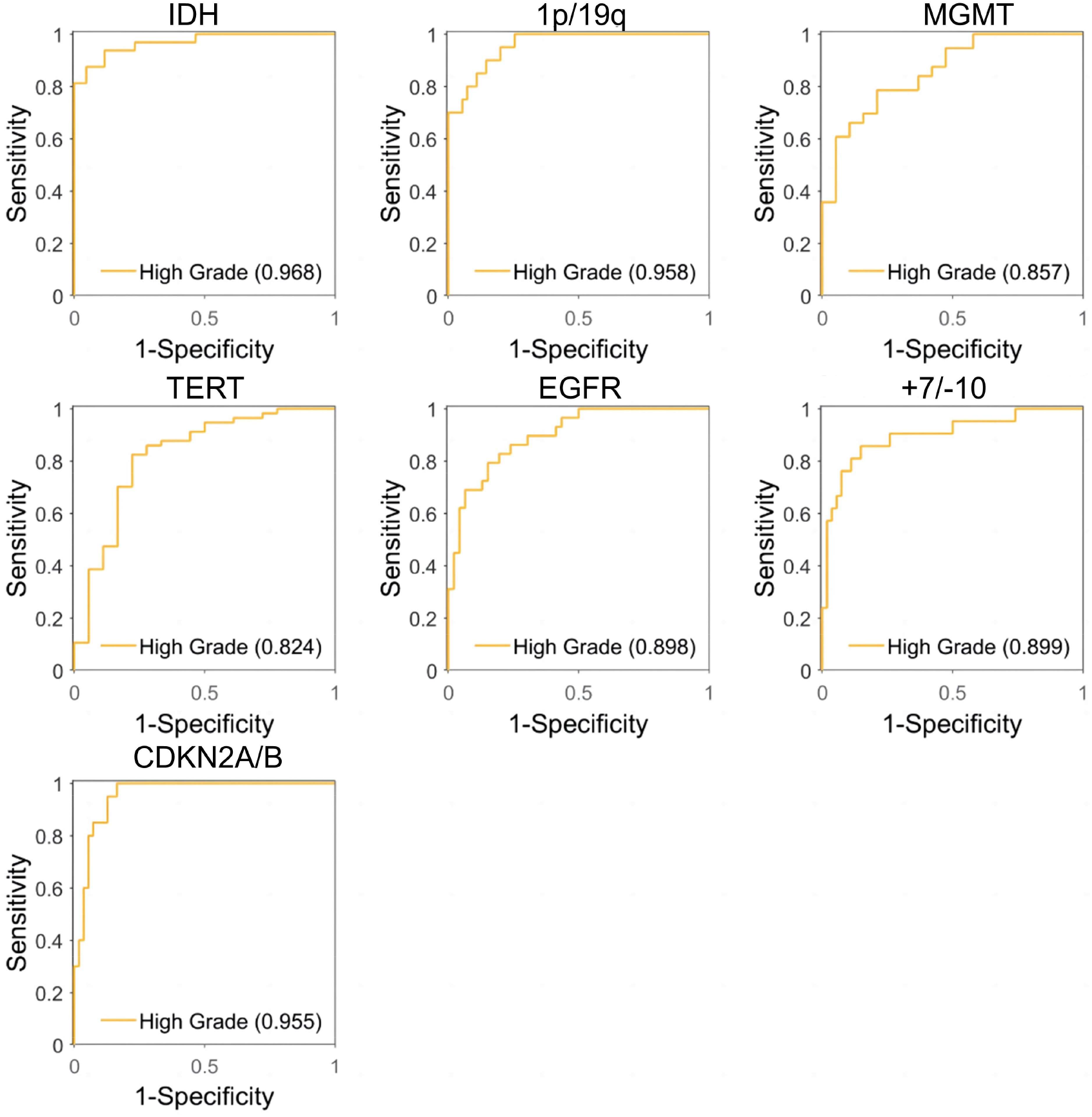
Comparisons of Raman diagnostic ROC using deep learning (ResNet) model. 7 molecular subgroups of high-grade glioma (**a**, IDH, **b**, 1p/19q, **c**, MGMT, **d**, TERT, **e**, EGFR, **f**, Chromosome7/10, **g**, CDKN2A/B) and 5 molecular subgroups of GBM (MGMT, TERT, EGFR, Chromosome7/10, CDKN2A and CDKN2B).

**Fig. 5.**
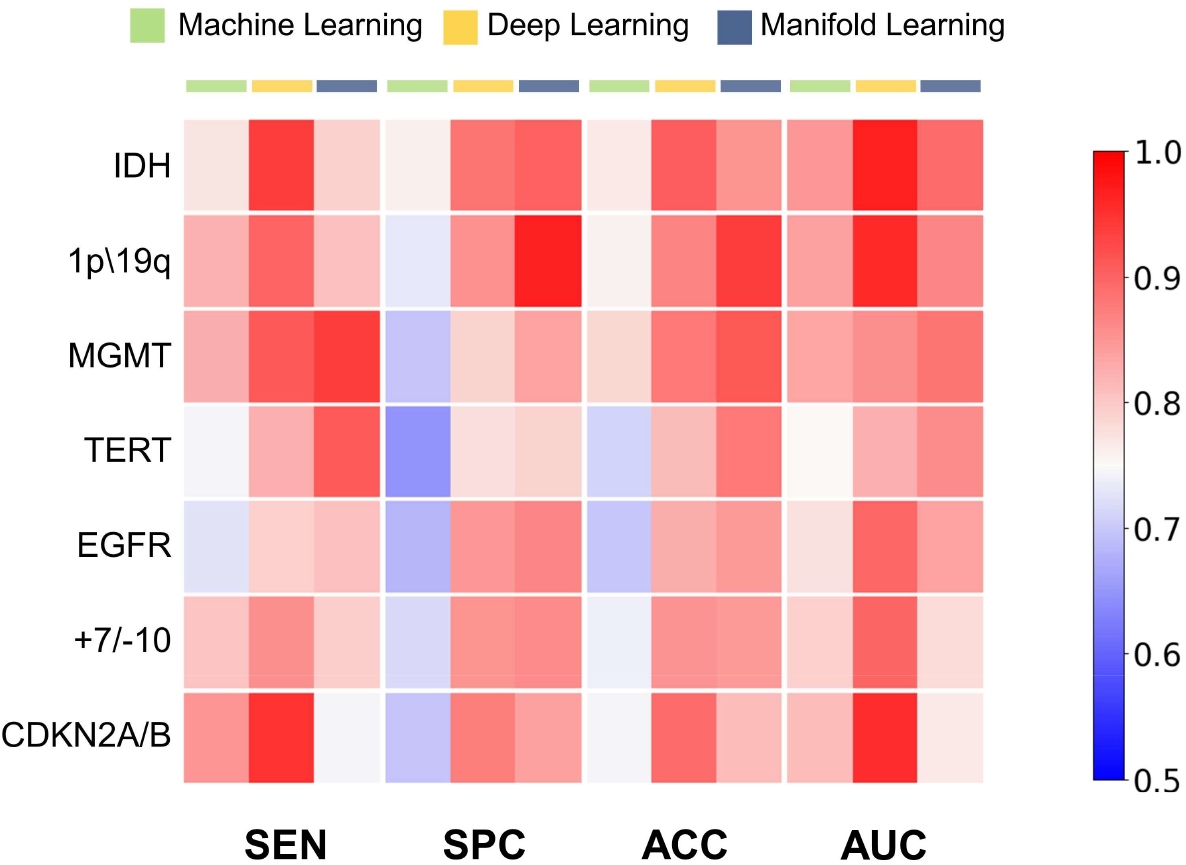
Heatmap of Raman diagnostic performance. For instance, sensitivity, specificity, accuracy and AUC of machine learning (SVM), manifold learning (UMAP), and deep learning (ResNet) models using Raman spectral datasets for separating high-grade glioma 7 molecular subgroups. SEN: sensitivity, SPC: specificity, ACC: accuracy, AUC: area under classification.

**Supplementary Figure S3-S6** compares the performance (sensitivity, specificity, accuracy and AUC) between machine learning (SVM), manifold learning (UMAP), and deep learning (ResNet), indicating that deep learning is a more practical algorithm in most subgroups. Comparing with PCA-SVM and UMAP in the **table S2**, ResNet was better for discriminating IDH mutation and 1p19q codeletion. Using ResNet also achieves better overall diagnostic accuracy in most of the other subgroups (82.67% in EGFR, 78.67% in Chromosome7/10 and 89.33% in CDKN2A/B). It is exceptional that in MGMT and TERT, using ResNet achieves lower diagnostic accuracy (88.00% in MGMT and 81.33% in TERT) than manifold learning (91.28% in MGMT and 87.92% in TERT).

### GBM molecular subgrouping with deep learning

To further demonstrate the effectiveness of our classification models, performance in GBM 5 molecular subgroups is also evaluated. In 422 tissue sites of 29 high-grade glioma patients, the Raman spectra are preprocessed, split and fed into the models in the same way. **Figure 6** shows the performance of subgrouping. With our deep learning model, the AUCs in all subgroups are larger than 0.85. The diagnostic accuracy in each subgroup is also better than 0.86 (over 0.9 in MGMT, TERT and CDKN2A/B), demonstrating our ResNet to be a practical algorithm for subgrouping.

**Fig. 6.**
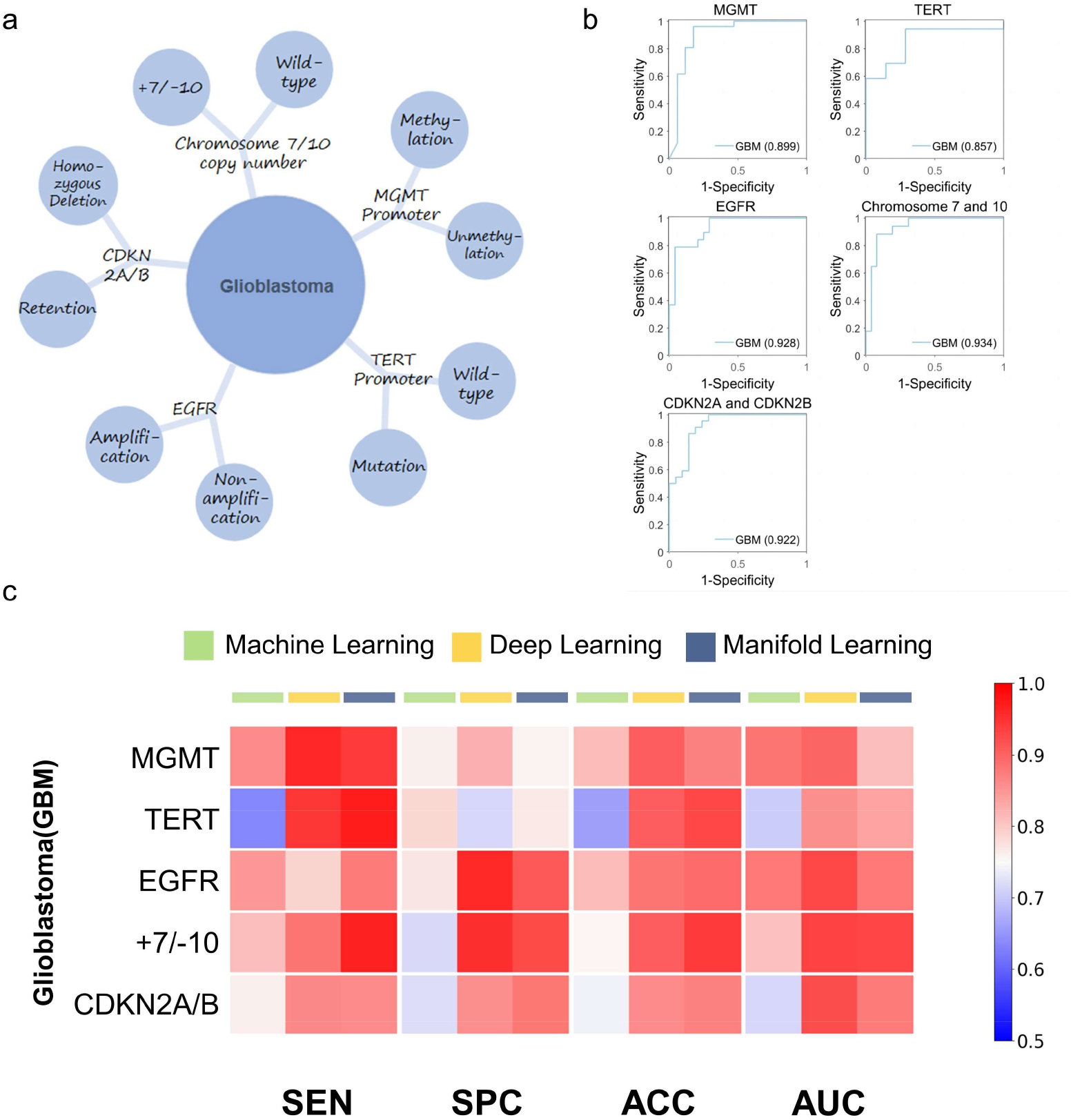
Classification performance in GBM 5 molecular subgroups. **a**, 5 typical molecular subgroups selected in GBM. **b**, Raman diagnostic ROC using deep learning (ResNet) model. **c**, Heatmap of Raman diagnostic sensitivity, specificity, accuracy and AUC of machine learning (SVM), manifold learning (UMAP), and deep learning (ResNet) models using Raman spectral datasets for separating GBM 5 molecular subgroups. SEN: sensitivity, SPC: specificity, ACC: accuracy, AUC: area under classification.

### Raman features related to high-grade glioma molecular subgrouping

To further investigate the significant biomolecular information identified in tissue Raman spectra during the glioma molecular diagnosis process of ResNet. Raw spectral region of interest (ROI) between molecular subgrouping, and ResNet determined spectral ROI which are most contributed to the diagnostic models are selected using the saliency maps, as shown in **Figure 7** and **table S3**. The Raman peaks related to the glioma tissues are recognized, reflecting the most significant biomolecular variance in Raman diagnosis based on the BSF method of ResNet. In IDH recognition, the saliency map was positive related to the Raman peaks of (499 cm^-1^, 568 cm^-1^, 577 cm^-1^ (phosphatidylinositol), 1078 cm^-1^ (lipid, phospholipid, nucleic acid), 1473 cm^-1^), negative related to the Raman peaks of (1448 cm^-1^ (collagen), 1433 cm^-1^ (lipid)). In 1p/19q recognition, the saliency map was positive related to the Raman peaks of (525 cm^-1^ (serine, cysteine), 547 cm^-1^ (cholesterol), 1085 cm^-1^ (nuclei acid), 1094 cm^-1^ (DNA)), negative related to the Raman peaks of (1319 cm^-1^ (collagen), 1332 cm^-1^ (CH^3^CH_2_ wagging, collagen)). In MGMT recognition, the saliency map was positive related to the Raman peaks of (930 cm^-1^ (collagen), 951 cm^-1^ (protein), 1560 cm^-1^ (tryptophan)), negative related to the Raman peaks of (640 cm^-1^ (tyrosine), 1442 cm^-1^ (triglycerides)). In TERT recognition, the saliency map was positive related to the Raman peaks of (529 cm^-1^), negative related to the Raman peaks of (508 cm^-1^, 525 cm^-1^, 536 cm^-1^ (cholesteryl esters), 540 cm^-1^ (cysteine)). In EGFR recognition, the saliency map was positive related to the Raman peaks of (1000 cm^-1^ (phenylalanine, collagen), 1221 cm^-1^ (protein)) negative related to the Raman peaks of (549 cm^-1^ (cholesterol), 850 cm^-1^ (tyrosine), 856 cm^-1^ (type I collagen)).

**Fig. 7.**
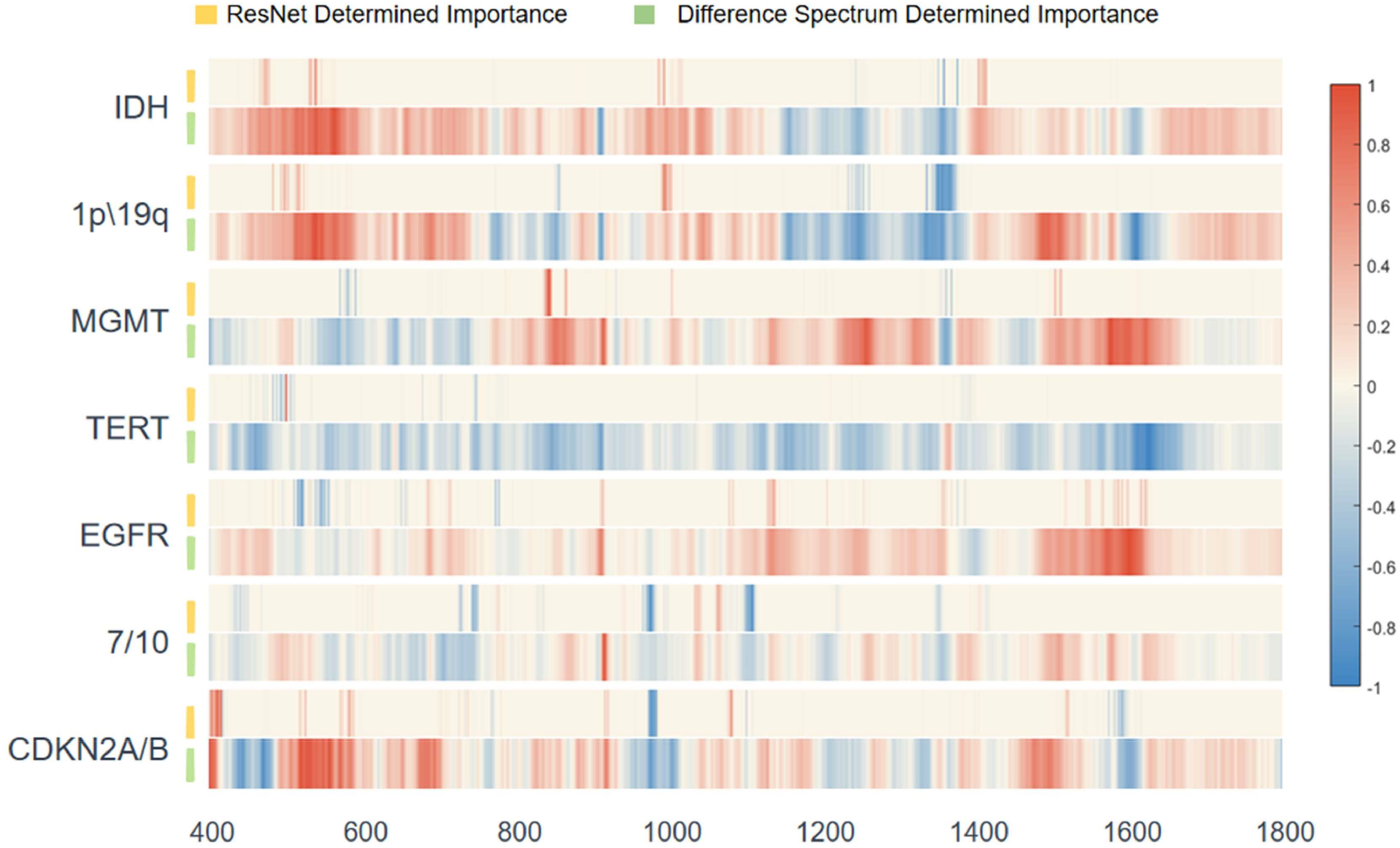
Saliency maps in ResNet model for Raman shift signatures. Discriminating high-grade glioma 7 molecular subgroups (IDH, 1p/19q, MGMT, TERT, EGFR, Chromosome7/10, CDKN2A/B) correlates the Raman shift of biomolecular subgroups, normalized with max values of difference spectrum.

In Chromosome copy number +7/-10 recognition, the saliency map was positive related to the Raman peaks of (1129 cm^-1^ (protein, lipid), 1155 cm^-1^ (protein, glycogen)), negative related to the Raman peaks of (818 cm^-1^ (collagen), 1068 cm^-1^(collagen, fatty acid, palmitic acid)). In CDKN2A/B recognition, the saliency map was positive related to the Raman peaks of (1169 cm^-1^ (type I collagen)), negative related to the Raman peaks of (1064 cm^-1^ (lipid), 1618 cm^-1^ (tryptophan), 1635 cm^-1^ (collagen)). The ResNet model recognized the relative intensity changes (either an increase or decrease) of multiple Raman bands related to biomolecules in the Raman spectra of high-grade glioma.

## Discussion

Distinguishing the molecular subgroups in the surgery is significant as it devotes to identify the operation plan. In this work, we have developed and validated the Raman diagnostic platform of the unique ResNet model for real-time accurate identification of molecular subgroups of high-grade glioma and GBM patients. The ResNet model was optimized with *tanh* activation function instead of *relu* or *linear* function. Nonlinear activation function achieves better fitting to many different spectroscopic patterns with better accuracy and less loss during training networks. The overall process including Raman spectra acquisition and model detection for single tissue site cost within 5 seconds. In AUC comparisons, ResNet was better for diagnosis than PCA-SVM and UMAP. All molecular subgrouping of high-grade glioma differentiates with the AUC of > 0.8 by using ResNet, especially IDH wild and mutation, 1p/19q intact and codeletion, CDKN2B depletion and no depletion differentiate with the AUC of >0.95. We also investigated GBM subgrouping to evaluate our Raman ResNet model. For the same molecular signature, of GBM subgrouping is better than high-grade glioma subgrouping in AUC comparisons, as shown in **Figure 6**.

For saliency map shown in **Figure 7**, the reduction of lipid and collagen reflects IDH wild. Cholesterol, serine, cysteine, and nuclei acid increase, reflecting 1p/19q intact. Tyrosine, protein classes, triglycerides, tryptophan and collagen change in glioma tissues, reflecting spectral difference between MGMT demethylation and methylation. Serine and cysteine decrease, reflecting TERT mutation. Cholesterol and tyrosine decrease, reflecting EGRF amplification. Collagen, lipid and porphyrin change, reflecting Chromosome7 amplification; proline, valine and phenylalanine change, reflecting Chromosome10 depletion. Phenylalanine and collagen change, reflecting CDKN2A depletion; tryptophan, lipid and collagen change, reflecting CDKN2B depletion. Previous studies have confirmed that the decline of lipids and collagen is closely related to IDH mutation. Mutated IDH subtyping would convert α-ketoglutaric acid in the tricarboxylic acid cycle into 2-hydroxy-glutaric acid (2-HG), and the high level of 2-HG inhibits the synthesis of triglycerides and other lipids^35^, and the high level of D2-HG blocks the prolyl hydroxylation of collagen, leading to defective maturation of collagen^36^. For other molecular signatures, literatures haven’t provided sufficient support.

Molecular classification could also have an immediate impact on the surgical strategies of patients with high-grade glioma. Surgical goals should be tailored based on molecular subgroups^37,38^. Patients with molecular astrocytoma who undergo gross total resection achieve a 5-year increase in median survival compared to patients who receive subtotal resections. Our Raman molecular subgrouping creates an avenue for accurate and rapid differentiation of high-grade glioma subgroups to define a better prognosis. Moreover, fewer than 10% of patients with glioma are enrolled in clinical trials^39^. Clinical trials limit inclusion criteria to a specific subpopulation, often defined by molecular subgroups. Our deep learning Raman method may offer a new opportunity for new clinical trials.

Current multiple techniques were developed for intraoperative brain cancer. Modalities such as ultrasound (US) and optical coherence tomography (OCT), confocal fluorescence microscopy (CFM) have been shown to provide structural information (large scale for US, sub-micrometer resolution for CFM, and microscopic scale for CFM, OCT) in real time. Intraoperative US show the ability to margin deep seated brain tumors from normal brain^40^. Intraoperative OCT has been able to distinguish high-density/low-density cancer in nine patients with high-grade gliomas^41^. Intraoperative confocal microscopy has shown evidence for invasion detection using fluorescence in grade 1 to 2 glioma on 10 patients^42^. However, no evidence indicates these modalities could investigate molecular subgroups. Recently, SRH has shown the possibility of detecting three molecular signatures (IDH, 1P/19q and ATRX) through stimulated Raman modality^33^, However, in practice, Raman mapping for molecular subtyping is not necessary with time consuming. Molecular subgrouping is on patient level and has no heterogeneity, thus, detection of several sites is more suitable for clinical situation, meanwhile, overall site-detection method is more cost-effective on both devices and algorithm.

This paper is a preliminary study confirmed that the feasibility of providing rapid molecular signature test to doctors during surgery. The influence of subdivided glioma categories on molecular feature recognition performance was discussed in this article, which can be applied in intraoperative scenarios to assist doctors in identifying GBM combined with rapid freezing pathological results. Our deep learning fiberoptic Raman spectroscopy may improve the detection performance of molecular features such as MGMT methylation etc. By incorporating with molecular weight analysis, we found that potential related makers (e.g. lipid and protein, DNA and collagen) for high-grade glioma subgrouping. Although we don’t implement in vivo molecular subgrouping detection, previous researches have shown the solid foundation of in vivo Raman spectroscopy^43,44^, indicating the feasibility of in vivo molecular subgrouping.

## Methods

### Tissue preparation

This study was approved by the ethical committee of Beijing Tiantan Hospital (KY2023-030-02). Before the operation, every patient underwent enhanced magnetic resonance imaging (MRI), including T1-weighted, T1-contrast, T2-weighted, and T2-Flair (T2-fluid-attenuated inversion recovery) modalities. Two experienced neuroradiologists independently reviewed the MRI data of patients, and those diagnosed with high-grade glioma (HGG) were considered for further analysis. During the operation, tumor tissues from patients diagnosed with HGG based on preoperative MRI were validated using rapid intra-operative tissue pathology. Only specimens showing tissue pathology consistent with the MRI images were collected. After the tumor tissue was resected, a sodium chloride solution was used to remove blood adhered to the tumor tissue. The original tumor tissue was then cut into several small tissue particles (approximately 2mm*2mm*2mm). Then the tumor samples were snap-frozen in liquid nitrogen and stored at -80 °C. All procedures were completed within 30 minutes. The tissues were sectioned and stained with hematoxylin-eosin (H&E) to confirm the pathological diagnosis of each sample. Finally, the tissues prepared on aluminum foil were used for Raman spectroscopic studies.

### Fiberoptic Raman spectroscopy

The Raman probe spectroscopy system as shown in **Supplementary Figure S1** which we used for high-grade glioma and GBM diagnosis, which is composed of Raman probe with filters (RamanProbe, Inphotonics Inc.), 785nm laser (o8NLDM, Cobolt Inc.) and high-sensitive spectrometer with ddpCCD (Acton 785, Princeton Instrumentation Inc.). The laser excitation power for the tissue Raman collection is 65mW, and the exposure time of single spectrum is 5 second. The numerical aperture (NA) of Raman probe (1cm in diameter) is 0.22.

### Molecular subgrouping status

This study included 7 astrocytoma, 8 oligodendroglioma, and 29 GBM (including 2 molecular GBM) from Beijing Tiantan Hospital. The enrolled patients’ data, including basic demographic information, imaging data, pathological diagnosis, and molecular characteristics, were collected from the hospital information system (HIS). The integrated diagnosis of tumors in this study relied on histological pathology and molecular features. The histological pathology of tumors was validated using the hematoxylin-eosin (H&E) stain of tumor tissue. The following key molecular features, which contribute to the integrated diagnosis of glioma, were determined using pyrosequencing and/or next-generation sequencing (NGS), including the mutation status of IDH1 and IDH2, MGMT promoter methylation status, 1p/19q co-deletion status, the mutation status of the TERT promoter, EGFR amplification status, gain of chromosome 7 and loss of chromosome 10, and the homozygous deletion status of CDKN2A/B. According to the 2021 WHO classification of tumors of the central nervous system, common diffuse gliomas in adults are divided into three types: astrocytoma, IDH-mutant; oligodendroglioma, IDH-mutant and 1p/19q co-deleted; and glioblastoma, IDH-wildtype. All IDH-mutant diffuse astrocytic tumors are considered as a single type (astrocytoma, IDH-mutant) and are graded as CNS WHO grade 2, 3, or 4. However, the presence of homozygous deletion of CDKN2A/B results in a CNS WHO grade of 4, even without typical histological features such as microvascular proliferation or necrosis. For IDH-wildtype diffuse gliomas in adults, if one or more of the three genetic parameters are present (TERT promoter mutation, EGFR gene amplification, combined gain of chromosome 7 and loss of chromosome 10), these lesions are assigned to the highest WHO grade, namely molecular GBM. Oligodendroglioma, IDH-mutant and 1p/19q co-deleted, are graded as CNS grade 2 or 3 based on histological features. Anaplastic oligodendroglioma is not included in this classification. Only in the context of an IDH-wildtype diffuse and astrocytic glioma in adults, if there are typical histological features (microvascular proliferation or necrosis) or key molecular characteristics (TERT promoter mutation or EGFR gene amplification or +7/-10 chromosome copy number changes), a diagnosis of glioblastoma, IDH-wildtype can be made.

### Raman spectrum pre-process

The original spectral data contains various noise and auto-fluorescence background; therefore, the spectra need to be processed before being input into the deep learning model. The pre-processing takes four steps: (1) wavenumber selection; (2) background subtraction; (3) smoothing; (4) normalization. In brief, the wavenumber between 400-1800 cm^-1^ was selected as the region of interest. The asymmetric least-squares method was applied to subtract the background signal. The data were then smoothed by a Savitzky-Golay filter to reduce the noise and increase the signal-to-noise ratio. All the processing mentioned above was done by Python 3.7 library scipy 1.8.0.

### Raman spectrum classification model

Using PCA, we assume that all meaningful information which contains within the variance. Through finding the maximum variance space, we could get principal components (PC1, PC2 etc.) of Raman shift with high variance. Using SVM, we create a hyper plane (ω*x-b=0) with minimum distance between points. SVM method only focuses on class weight from extreme points, in the meanwhile ignoring distances between other points to hyper plane. Therefore, SVM may fit well with small sample size Raman spectrum data. Combing PCA and SVM, we built a machine learning based classification model for high-grade glioma subgrouping. All the processing mentioned above was done by Python 3.7 library sklearn 0.24.2.

Different with PCA, UMAP used nonlinear dimensional reduction, and find a representation (UMAP1, UMAP2 etc.) of Raman data in low-dimensional space R^N^. Firstly, a good map from Riemannian manifold M to R^N^ was found. Then Raman data D is uniformly drawn from M. By simulating approximate distances in M between points in D that are close enough in R^N^, we finally get UMAP values in R^N^. Here, we built supervised UMAP classification model for high-grade glioma subgrouping. All the processing mentioned above was done by Python 3.7 library UMAP-learn 0.5.3.

Structure of ResNet were described in the **Figure 2B**. Here we optimized the 1D ResNet networks for Raman data modeling using following strategies. (1) In order to achieve the capture of information at different scales, multi-layer convolution operations are used. (2) To simulate many different data patterns, nonlinear activation functions Tanh are used instead of *Linear* or *Relu*. (3) To achieve accurate optimization of hyper parameters of network, backpropagation and gradient descent are used to fit spectral wavenumber information. (4) The saliency map of deep learning models (ResNet) was simulated by the binary stochastic filtering (BSF) feature selection methods as shown in **Figure 2B**. All these optimization steps efficiently extract one-dimensional Raman spectral data information for binary subgrouping, and avoid model over-fitting. All the processing mentioned above was done by Python 3.7 Library keras 2.2.4 and tensorflow 1.14.0.

### Model evaluation

For subgrouping evaluation, true positives correspond to correct molecular subgrouping of Raman spectra from each tissue site compared with the ground truth of Glioma from the same tissue, false positives correspond to wrong subgrouping for instance (e.g., IDH wild), and false negatives correspond to wrong subgrouping for instance (e.g., IDH mutation). Outcomes of high-grade glioma Raman classification were evaluated with respect to sensitivity (SEN), specificity (SPC), and accuracy (ACC) as follows:

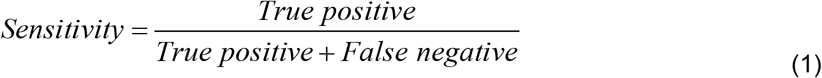

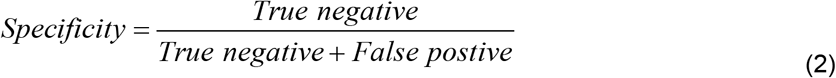

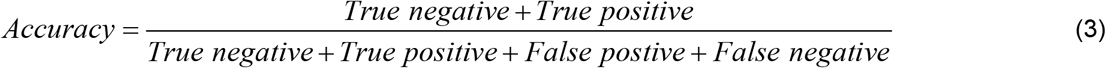

The binary classification for glioma molecular subgrouping was evaluated with a binary receiver operating characteristic (ROC) analysis according to the method. A ROC curve was generated by continuously varying the threshold of the probability for each category based on the ground truth. The area under the ROC curve (AUC) ranging from 0 to 1 evaluates the ability of a model to accurately distinguish different glioma subgroups with maximum sensitivity and specificity.

## Data Availability

All data produced in the present study are available upon reasonable request to the authors.

## Data availability and Code availability

The main data supporting the findings of this study are available within the paper and its Supplementary Information. All the programs for research purposes are available upon reasonable request. The system control software and the data collection software are proprietary and used in licensed technologies.

### Acknowledgements

This work was supported by National Natural Science Foundation of China (No. 62027824, No. 91959120, and No. 62205010); Beijing Natural Science Foundation (No.7224367 and No. L223018); Fundamental Research Funds for the Central Universities (No. YWF-22-L-547 and No. YWF-22-L-1265).

## Author contributions

C.L. and J.W. contribute equally. X.C., N.J. and S.Y. designed the methodology. J.W. and N. J. provided glioma tumor specimens and RNA sequencing results. C.L. and J.W. performed experiments using Raman spectroscopy. C.L. and J.S. wrote data processing programs. X.C., S. Y. and N.J. supervised the project., C.L., N.J., X. C. and S. Y. wrote the manuscript. All authors edited the manuscript.

## Competing interests

The authors declare no competing interests.

## Notes

### Competing Interest Statement

The authors have declared no competing interest.

### Author Declarations

Ethics committee/IRB of Beijing Tiantan Hospital and Beihang University gave ethical approval for this work.

