## Supporting information for "Accurate and Rapid Molecular Subgrouping of High-Grade Glioma via Deep-learning-assisted Label-free Fiberoptic Raman Spectroscopy"

**Supplementary Materials**

Fig. S1 Setup of fiberoptic Raman spectroscopy.

Fig. S2 Mean FP Raman spectra of GBM molecular subtyping for classification acquired from 422 tissue sites of 27 patients under Raman spectroscopy.

Fig. S3 Raman diagnostic ROC using manifold learning (UMAP) model for 7 molecular subgroups of high-grade glioma.

Fig. S4 Raman diagnostic ROC using machine learning (PCA-SVM) model for 7 molecular subgroups of high-grade glioma.

Fig. S5 Raman diagnostic ROC using manifold learning (UMAP) model for 5 molecular subgroups of GBM.

Fig. S6 Raman diagnostic ROC using machine learning (PCA-SVM) model for 5 molecular subgroups of GBM.

Table S1 Patient demographics and glioma subtyping diagnosis.

Table S2 Performance of molecular subtyping by ResNet model.

Table S3 Raman peaks of each molecular subtype of ResNet on saliency map.

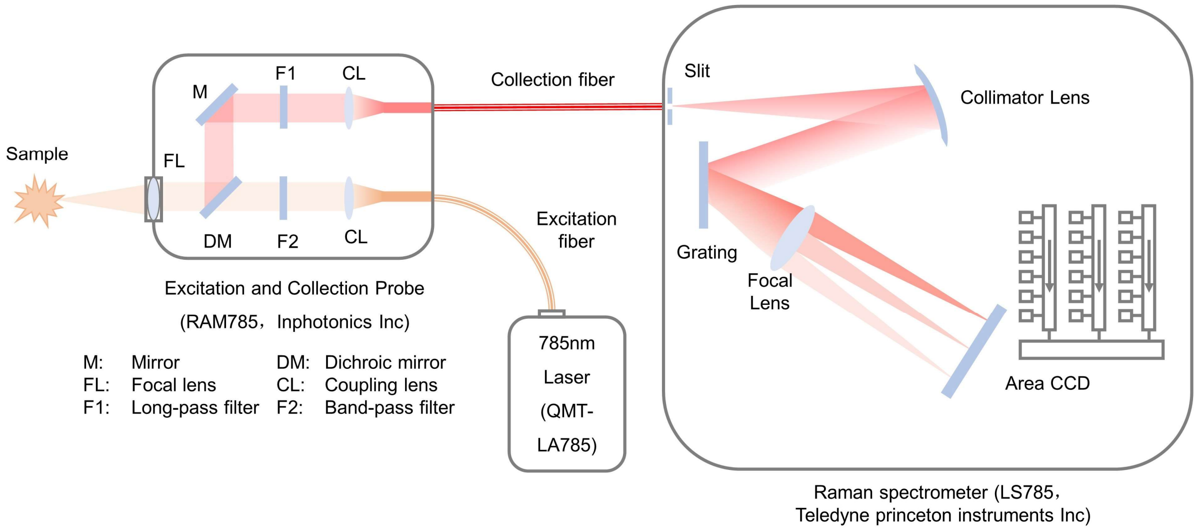


**Fig. S1 | Setup of fiberoptic Raman spectroscopy.**

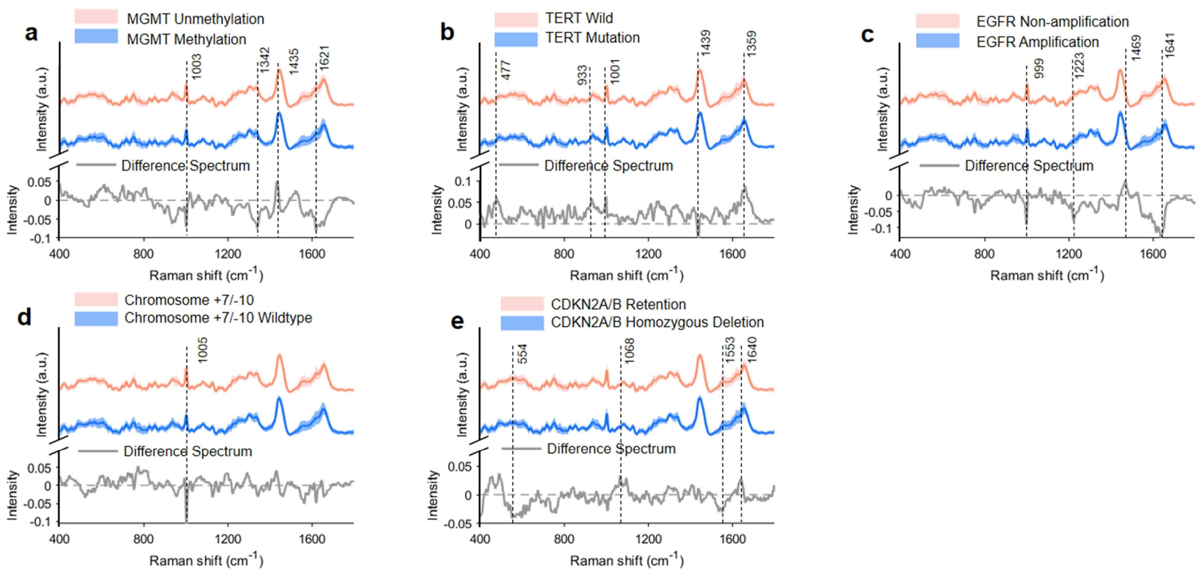


**Fig. S2 | Mean FP Raman spectra of GBM molecular subtyping for classification acquired from 422 tissue sites of 27 patients under Raman spectroscopy. a**, MGMT unmethylation and methylation tissues (methylation: 546; unmethylation: 197). **b**, TERT wildtype and mutation tissues (mutation: 508; wildtype: 235). **c**, EGFR non-amplification and amplification tissues (amplification: 241; non-amplification: 502). **d**, Chromosome7 non-amplification and amplification tissues (amplification: 322; non-amplification: 421). **e**, Chromosome10 non-deletion and deletion tissues (deletion: 242; non-deletion: 501). **f,** CDKN2A non-deletion and deletion tissues (deletion: 276; non-deletion: 467). **g,** CDKN2B non-deletion and deletion tissues (deletion: 226; non-deletion: 517).

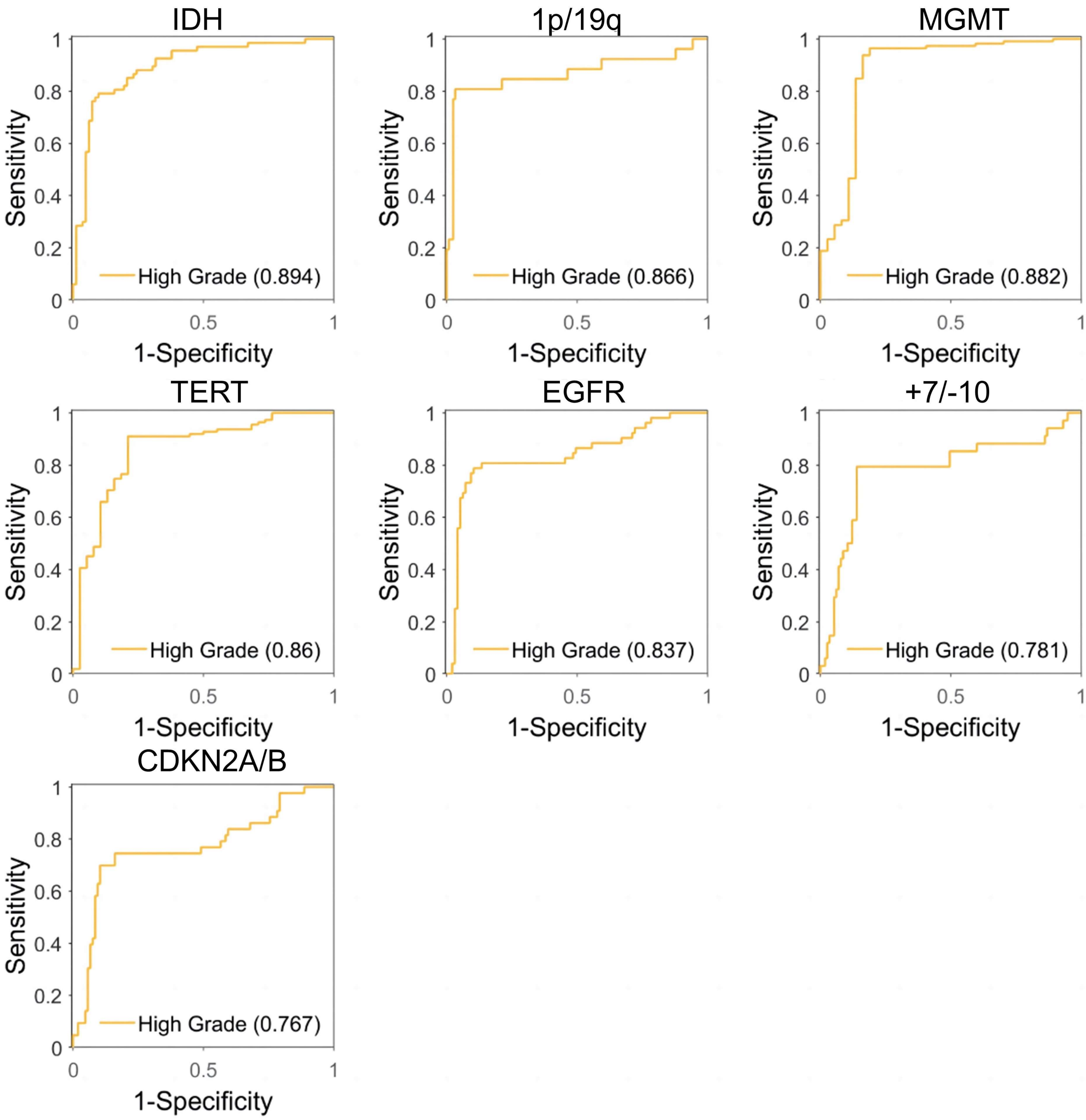


**Fig. S3 | Raman diagnostic ROC using manifold learning (UMAP) model.** 7 molecular subtypes of high-grade glioma (IDH, 1p/19q, MGMT, TERT, EGFR, Chromosome7/10, CDKN2A/B).

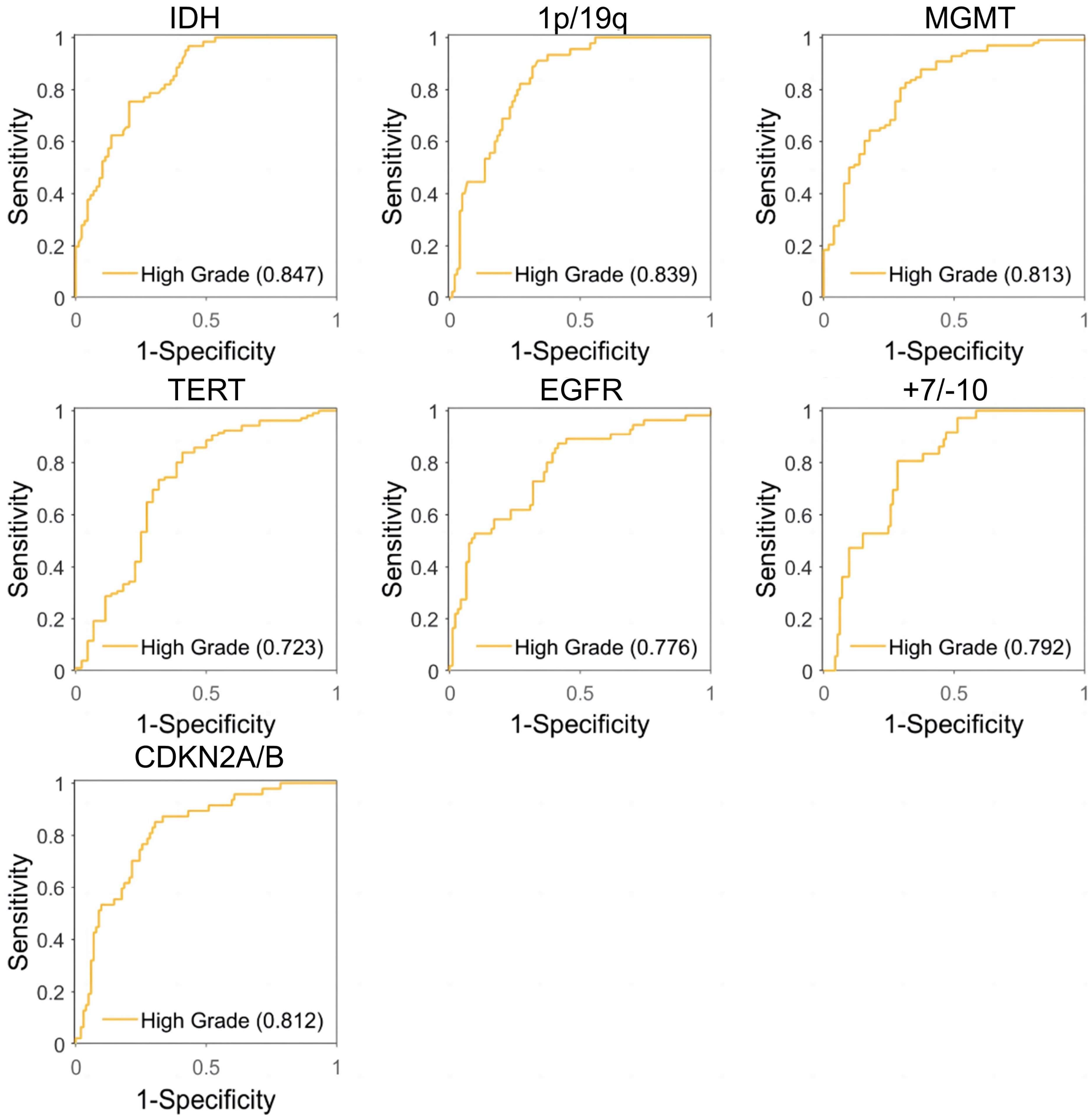


**Fig. S4 | Raman diagnostic ROC using machine learning (PCA-SVM) model.** 7 molecular subtypes of high-grade glioma (IDH, 1p/19q, MGMT, TERT, EGFR, Chromosome7/10, CDKN2A/B).

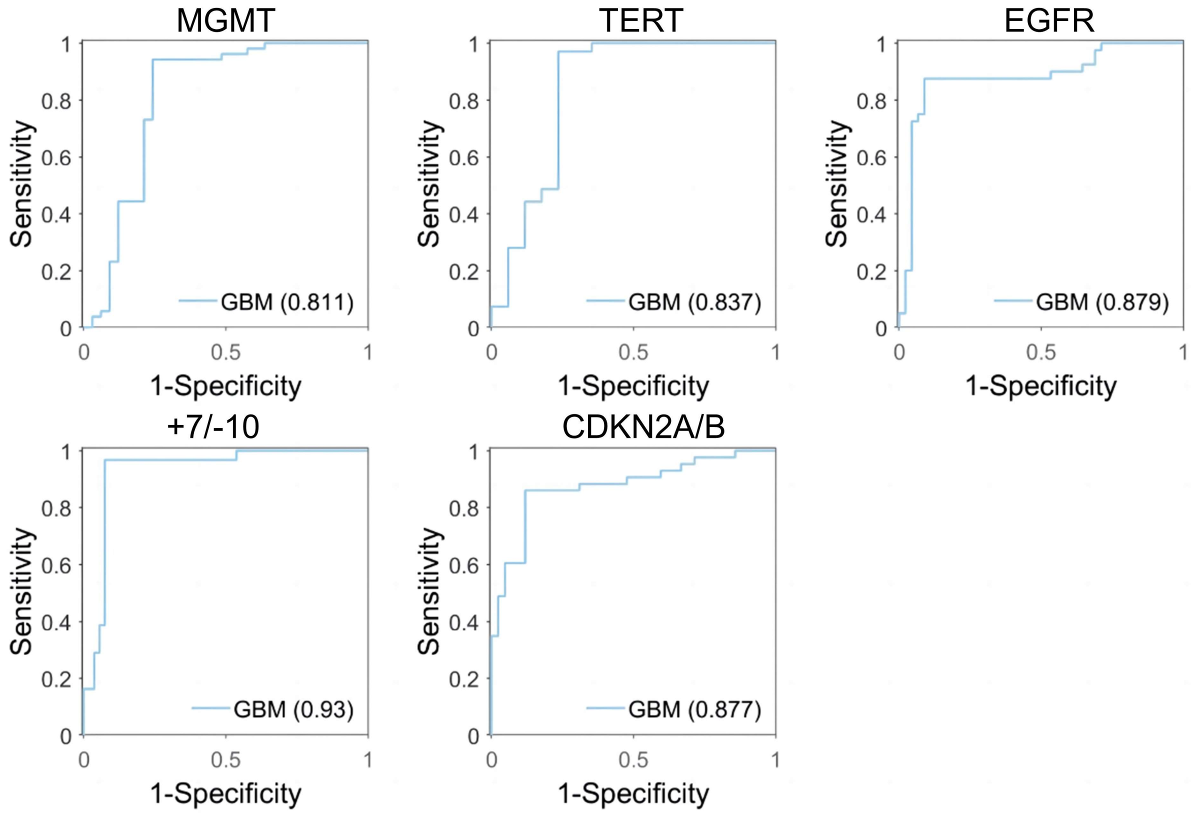


**Fig. S5 | Raman diagnostic ROC using machine learning (UMAP) model.** 5 molecular subtypes of high-grade glioma (MGMT, TERT, EGFR, Chromosome7/10, CDKN2A/B).

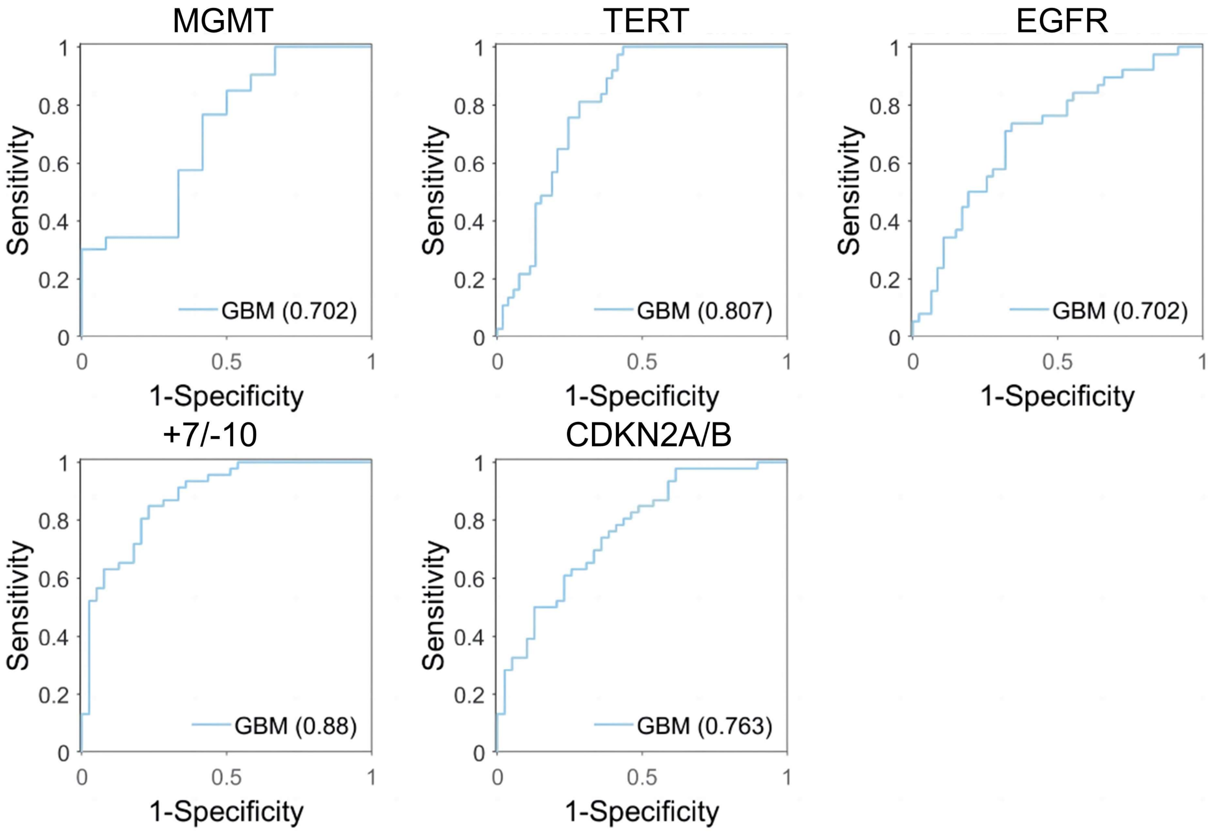


**Fig. S6 | Raman diagnostic ROC using machine learning (PCA-SVM) model.** 5 molecular subtypes of high-grade glioma (MGMT, TERT, EGFR, Chromosome7/10, CDKN2A/B).

**Tables**

**Table S1 | Patient demographics and glioma subtyping diagnosis.**

| Grade | Patients | Tissues | Spectrum | Spectrum | | | |
|---|---|---|---|---|---|---|---|
| | | | | IDH wild | 1p/19q intact | MGMT[1] | TERT[2] |
| GBM (percentage) | 27 | 61 | 422 | 422 | 422 | 235 | 348 |
| | | | | | | 55.69% | 82.46% |
| Molecular GBM | 2 | 2 | 20 | 20 | 20 | 10 | 20 |
| astrocytoma | 7 | 15 | 151 | 0 | 151 | 151 | 0 |
| oligodendroglioma | 8 | 14 | 150 | 0 | 0 | 150 | 140 |
| total | 44 | 92 | 743 | 59.49% | 79.81% | 73.49% | 68.37% |

| Grade | Patients | Tissues | Spectrum | Spectrum | | |
|---|---|---|---|---|---|---|
| | | | | EGFR[3] | Chromosome7/10[4] | CDKN2A/B[5] |
| GBM (percentage) | 27 | 61 | 422 | 201 | 182 | 206 |
| | | | | 47.63% | 43.13% | 48.82% |
| Molecular GBM | 2 | 2 | 20 | 10 | 10 | 20 |
| astrocytoma | 7 | 15 | 151 | 0 | 0 | 0 |
| oligodendroglioma | 8 | 14 | 150 | 30 | 0 | 0 |
| total | 44 | 92 | 743 | 32.44% | 25.84% | 30.42% |

1: MGMT methylation, 2: TERT wildtype, 3: EGFR amplification, 4: Gain of entire Chromosome 7 and Loss of entire Chromosome 10, 6: CDKN2A/B retention.

**Table S2 | Performance of molecular subtyping by ResNet model.**

| Molecular subtyping | SEN[1] | SPC[2] | ACC[3] | AUC[4] | Time(s) | SEN | SPC | ACC | AUC | Time(s) |
|---|---|---|---|---|---|---|---|---|---|---|
| IDH | 93.75% | 88.37% | 90.67% | 0.968 | 10.0 | / | / | / | / | / |
| 1p/19q | 90.00% | 85.45% | 86.67% | 0.958 | 10.0 | / | / | / | / | / |
| MGMT | 91.07% | 78.95% | 88.00% | 0.857 | 9.8 | 96.15% | 82.35% | 90.70% | 0.899 | 10.4 |
| TERT | 82.46% | 77.78% | 81.33% | 0.824 | 12.0 | 94.44% | 71.43% | 90.70% | 0.857 | 10.4 |
| EGFR | 79.31% | 84.78% | 82.67% | 0.898 | 10.5 | 78.95% | 95.83% | 88.37% | 0.928 | 11.5 |
| +7/-10 | 85.71% | 85.19% | 85.33% | 0.899 | 10.6 | 88.24% | 92.31% | 90.70% | 0.934 | 9.97 |
| CDKN2A/B | 95.00% | 87.27% | 89.33% | 0.955 | 11.0 | 86.36% | 85.71% | 86.05% | 0.922 | 10.5 |

1: sensitivity, 2: specificity, 3: accuracy, AUC: area under curve.

**Table S3 | Raman peaks of each molecular subtype of ResNet on saliency map.**

| Molecular subtyping | Raman shift($cm^{-1}$) | Weight | Related molecules |
|---|---|---|---|
| IDH | 499 | 0.31 | / |
| | 568 | 0.49 | / |
| | 577 | 0.53 | phosphatidylinositol |
| | 1078 | 0.62 | lipid, phospholipid, nucleic acid |
| | 1433 | -0.32 | lipid |
| | 1448 | -0.44 | collagen |
| | 1473 | 0.49 | / |
| 1p/19q | 525 | 0.73 | serine, cysteine |
| | 547 | 0.6 | cholesterol |
| | 1085 | 0.96 | nuclei acid |
| | 1094 | 0.46 | DNA |
| | 1319 | -0.31 | collagen |
| | 1332 | -0.1 | $CH_3CH_2$ wagging, collagen |
| MGMT | 640 | -0.63 | tyrosine |
| | 930 | 0.6 | collagen |
| | 951 | 0.63 | protein |
| | 1442 | -0.5 | triglycerides |
| | 1560 | 0.51 | tryptophan |
| TERT | 508 | -0.36 | / |
| | 525 | -0.44 | / |
| | 529 | 1 | / |
| | 536 | -0.44 | cholesteryl esters |
| | 540 | -0.32 | cysteine |

| Molecular subtyping | Raman shift($cm^{-1}$) | Weight | Related molecules |
|---|---|---|---|
| EGFR | 549 | -0.58 | cholesterol |
| | 850 | -0.49 | tyrosine |
| | 856 | -0.3 | type I collagen |
| | 1000 | 0.6 | phenylalanine, collagen |
| | 1221 | 0.46 | protein |
| +7/-10 | 449 | −0. 43 | / |
| | 818 | −0. 81 | collagen |
| | 1068 | −0. 99 | collagen, fatty acid, palmitic acid |
| | 1129 | 0. 64 | protein, lipid |
| | 1155 | 0. 48 | protein, glycogen |
| | 1196 | −1 | / |
| | 1427 | −0. 3 | / |
| CDKN2A/B | 1064 | -0.66 | lipid |
| | 1169 | 0.35 | type I collagen |
| | 1618 | -0.33 | tryptophan |
| | 1635 | -0.44 | collagen |
